# Neural correlates of aggression outcome expectation and their association with aggression: A voxel-based morphometry study

**DOI:** 10.1101/2023.08.26.23294598

**Authors:** Xinyu Gong, Bohua Hu, Liang Wang, Qinghua He, Ling-Xiang Xia

## Abstract

**Background:** The aggression outcome expectation is believed to be an important cognitively influencing factor of aggression. Discovering the neural mechanism of aggression outcome expectation is conductive to develop aggression research. However, the neural correlates underlying aggression outcome expectation and its deleterious effect remain elusive.

**Methods:** In this study, we utilized voxel-based morphometry (VBM) to unravel the neural architecture of aggression outcome expectation measured by the Social Emotional Information Processing Assessment for Adults and and its relationship with aggression measured by the Buss Perry Aggression Questionnaire in a sample of 185 university students (114 female; mean age = 19.94 ± 1.62 years; age range: 17-32 years).

**Results:** The results showed a significantly positive correlation between aggression outcome expectation and the regional gray matter volume (GMV) in the right middle temporal gyrus (MTG) (x = 55.5, y = -58.5, z = 1.5; t =3.35; cluster sizes =352, *p* < 0.05, GRF corrected). Moreover, aggression outcome expectation acted as a mediator underlying the association between the right MTG volume and aggression.

**Conclusions:** These results found the neural correlates of aggression outcome expectation and its effect on aggression for the first time, which may contribute to our understanding of the cognitive neural mechanism of aggression and tentatively provide an effective method to identify potential neurobiological markers for aggression.

## Introduction

Aggression outcome expectation refers to the cognitive response or inclination individuals have towards anticipating the potential outcomes resulting from aggressive actions (Arsenio et al., 2009; Coccaro et al., 2017; Crick & Dodge, 1996; Fernández-Fuertes et al., 2019). The Social Information Processing Model posits that aggression outcome expectation is an important cognitive motivator of aggression (Crick & Dodge, 1994; Crick & Dodge, 1996). And the main function of aggression outcome expectation is leading to aggressive behavior. Thus, unravel the neural mechanism underlying aggression outcome expectation holds significant significance in understanding mechanism of aggressive cognition and behavior (Hubbard et al., 2001; Lischinsky & Lin, 2020; Marsee & Frick, 2007; Wei et al., 2023) and identification of potential neurobiological markers for aggression. Furthermore, it can aid in the development of targeted interventions and preventive measures to mitigate aggressive cognition and behavior from the perspective of the brain (Allen et al., 2018; Anderson & Bushman, 2002; Fritz et al., 2023; Li et al., 2022; Smithmyer et al., 2000). However, the neural basis of aggression outcome expectation and its association with aggressive behavior remain understudied. In this study, our primary objective was to investigate the neural substrates supporting aggression outcome expectation and its effect on aggression in a sample of university students.

Voxel-Based Morphometry (VBM), which involves analyzing structural brain imaging data to examine variations in gray matter density or volume across individuals or groups, offers valuable insights into the biological underpinnings of these psychological aspects (Pan et al., 2023). It was a widely used technique that provided valuable insights into the neural architecture associated with psychological variable (Blum et al., 2023; Wang et al., 2023; Zhang et al., 2023). This technique plays a significant role in unraveling the neural foundations of aggression cognition and personality traits, thus illuminating the intricate relationship between neural structure and psychological traits (Ide et al., 2020; Wei et al., 2023). Hence, we conducted a voxel-based morphometry (VBM) study to investigate the neural correlates of aggression outcome expectation. By applying VBM to structural brain imaging data, we aimed to identify specific brain regions where regional gray matter volume (GMV) associated with aggression outcome expectation.

Since the neural correlates of aggression outcome expectation have not been reported, we could only infer brain-related implications from the psychological components and essential attributes of aggression outcome expectation, the neural correlates of which have been explored. Firstly, aggression outcome expectation refers to the ability of conditional reasoning regarding the relationship between aggressive actions and their potential consequences, representing a form of consequential thinking (Guerra & Slaby, 1989), which involves the frontal lobes, particularly the prefrontal cortex (Dumontheil, 2014). Secondly, anticipatory imagination, including anticipation of aggression-related rewards, is involved in aggression outcome expectation (Kruithof et al., 2023). Anticipatory imagination is linked to brain regions associated with mental imagery and prospective thinking, which are often associated with the lateral and ventromedial prefrontal cortex (Diekhof et al., 2011). Thirdly, in terms of its fundamental attributes, aggression outcome expectation reflects an instrumental motivation for aggression (Smithmyer et al., 2000). Instrumental motivation involves brain circuits related to reward processing and goal-directed behavior, which are found in the frontal and temporal lobes, such as the medial prefrontal cortex (MPFC), inferior frontal gyrus (IFG) and middle temporal gyrus (MTG)(Zhu et al., 2022). At last, extensive research have indicated that reward anticipation mainly involve activations in the MPFC and IFG(Chen et al., 2022; Gorka et al., 2015; Yang et al., 2022). Given that brain structures primarily associated with these factors are located in the frontal and temporal lobes (Barkley-Levenson & Galván, 2014; Coccaro et al., 2018; Denson et al., 2009; Hajcak et al., 2005; Mok et al., 2009), we hypothesized that the neural basis of aggression outcome expectation predominantly focused on the frontal and temporal volumes.

Moreover, structural alterations in brain regions, such as variations in gray matter volume within the prefrontal cortex and temporal lobes, may also be linked to aggressive behavior (Gong et al., 2022; Holz et al., 2023; Zhu et al., 2019). It could be inferred that the effect of aggression outcome expectation on aggression may also have neuroanatomical bases. We hypothesized that aggression outcome expectation would mediate the relationship between the frontal and temporal volumes and aggression.

## Methods

### Participants

A total of 185 university students (114 female; mean age = 19.94 ± 1.62 years; age range: 17-32 years) were recruited from Southwest University in China for this study. All participants had no prior history of mental or neurological disorders and were right-handed. After signing the informed consent form, participants completed a scenario questionnaire assessing aggression outcome expectation, an aggression questionnaire, and underwent magnetic resonance imaging (MRI) scans. The study was approved by the Research Project Ethical Review Committee of the Faculty of Psychology at Southwest University, China. All participants were compensated for their participation. To ensure the accuracy of the results, participants were screened based on the quality of the MRI images; one participant was excluded due to anatomical abnormality, and three participants were excluded due to an IQR (image quality rating) below C+ (Gao et al., 2022).

### Measurements

#### Aggression outcome expectation

The aggression outcome expectation was measured by the aggression outcome expectation subscale within the Social Emotional Information Processing Assessment for Adults (SEIP-Q; Coccaro et al., 2017). This questionnaire consisted of 8 conflict-laden vignettes that prompted participants to envision themselves as the protagonists in these scenarios. In each vignette, participants responded to 16 questions related to their expectation of the outcomes resulting from their aggressive behavior (For example, “If you attacked, how much would this person respect you?”). The aggression outcome expectation was calculated as the average of responses to the 128 questions, with higher scores indicating a stronger expectation of favorable outcomes resulting from engaging in aggressive behaviors. The Chinese version of the questionnaire demonstrated good reliability and validity in China (Quan et al., 2019; Wang et al., 2018). In this study, this scale demonstrated high internal consistency, with a Cronbach’s alpha coefficient of 0.95.

#### Aggression

The Buss-Perry Aggression Questionnaire (BPAQ; Buss & Perry, 1992) was a widely used and well-established self-report measure designed to assess aggressive behavior. The BPAQ consisted of 29 items (e.g., “I have trouble controlling my anger.”), each rated on a 5-point Likert scale, ranging from 1 (extremely uncharacteristic of me) to 5 (extremely characteristic of me). To compute the BPAQ total score, the responses from all items were summed up. Higher scores on the BPAQ indicated a greater tendency towards aggressive behavior. The Chinese version of the questionnaire demonstrated good reliability and validity in China (Wang et al., 2023; Li et al., 2022). In this study, this scale demonstrated high internal consistency, with a Cronbach’s alpha coefficient of 0.91.

### Data acquisition

The 3T Siemens Trio MRI scanner (Siemens Medical, Erlangen, Germany) was used to collect brain imaging data at the Brain Imaging Center, Southwest University. The MP-RAGE (magnetization-prepared rapid gradient echo) sequence was employed to acquire high-resolution T1 images with the following parameters (Wang et al., 2022; Zhu et al., 2019): repetition time/echo time/inversion time = 1900/2.52/900 ms, flip angle = 9°, acquisition matrix = 256 × 256, voxel size = 1 × 1 × 1 mm^3^, and number of slices = 176.

### Image preprocessing

All data were preprocessed using CAT12 toolbox (The Computational Anatomy Toolbox, https://neuro-jena.github.io/cat/, version 12.8 (r2159)), which integrates with SPM12 software (Statistical Parametric Mapping, https://www.fil.ion.ucl.ac.uk/spm/software/spm12/, version 7771) based on Matlab R2018b. This toolbox has been proved to be reliable in Voxel-Based Morphometry (VBM; Fillmer et al., 2018).

Preprocessing is carried out according to the CAT12 manual (https://neuro-jena.github.io/cat12-help/): T1 images were automatically segmented with reference to the TPMs into gray matter (GM), white matter (WM) and CSF. They were then affine registered to a MNI template using the Diffeomorphic Anatomical Registration Through Exponentiated Lie Algebra (DARTEL) tools (Ashburner, 2007). The output GM images were smoothed with the recommended 8 mm Full Width at Half Maximum (FWHM) Gaussian kernel. In addition, participants’ total intracranial volume (TIV) was calculated to serve as a covariable in subsequent statistical analysis.

### Statistical Analysis

In order to identify the brain regions associated with aggression outcome expectation, we conducted an exploratory whole-brain multiple regression analysis using voxel-based morphometry (VBM). We included aggression outcome expectation as the variable of interest, and age, gender, and total intracranial volume (TIV) as covariates of no interest. To avoid edge effects, an absolute threshold masking of 0.1 was applied. Gaussian Random Field theory (GRF) was used for multiple comparison correction, with a statistical threshold of voxel *p* < 0.001 and cluster *p* < 0.05, using the Data Processing & Analysis for Brain Imaging (DPABI) toolbox (V7.0_230110; Yan et al., 2016).

To explore the relationship between brain structures, aggression outcome expectation and aggression, we used the PROCESSv3.0 macro of SPSS to conduct mediation analyses (Hayes, 2017). We tested two models: one with brain structure as the independent variable, aggression as the dependent variable, and aggression outcome expectation as the mediator variable, with age, gender and TIV as control variables; and the other with brain structure as the mediator variable to examine whether brain structures mediate the influence of aggression outcome expectation on aggression.

### Confirmatory Cross-Validation

We used a four-fold cross-validation based on machine learning algorithms and linear regression to assess the stability of the relationship between aggression outcome expectation and brain structures (Kong et al., 2020). Grey matter volume was used as the independent variable and aggression outcome expectation as the dependent variable. The data was divided into four folds to ensure the balance of the distributions of the independent and dependent variables. Three of the folds were used to build a linear regression model, which was then applied to the fourth fold to obtain the predicted value. This process was repeated four times and the average of the results was calculated to obtain the r (predicted, observed) (i.e., the correlation of the observed values with the predicted values). Finally, a nonparametric testing method was used to test the significance of the model by generating 1,000 surrogate data sets.

## Results

### Behavioral results

Table 1 lists descriptive statistics of all variables and TIV (including mean, standard deviation, range, skewness, and kurtosis). TIV was not significantly correlated with aggression outcome expectation, *r* = 0.029, *p* > 0.05; and aggression, *r* = 0.09, *p* > 0.05. aggression outcome expectation and aggression both follow the normal distribution, *p* > 0.05, Kolmogorov-Smirnov test (Figure 1A), a significant positive correlation was observed between them after controlling for the effects of age, gender, and TIV, *r* = 0.38, *p* < 0.001 (Figure 1B).

**Table 1.** Descriptive statistics of All Variables (N = 185)

| Variables | Mean | SD | Range | Skewness | Kurtosis |
| --- | --- | --- | --- | --- | --- |
| Age (age) | 19.94 | 1.62 | 17-32 | 3.05 | 18.38 |
| TIV (mm<sup>3</sup>) | 1500.45 | 135.34 | 1206.31-1876.13 | 0.26 | -0.36 |
| aggression outcome expectation | 0.97 | 0.36 | 0.08-2.09 | 0.22 | 0.46 |
| Aggression | 2.20 | 0.56 | 1.14-3.90 | 0.47 | -0.20 |

N = number, SD = Standard Deviation, TIV: Total Intracranial Volume.

**Figure 1.**
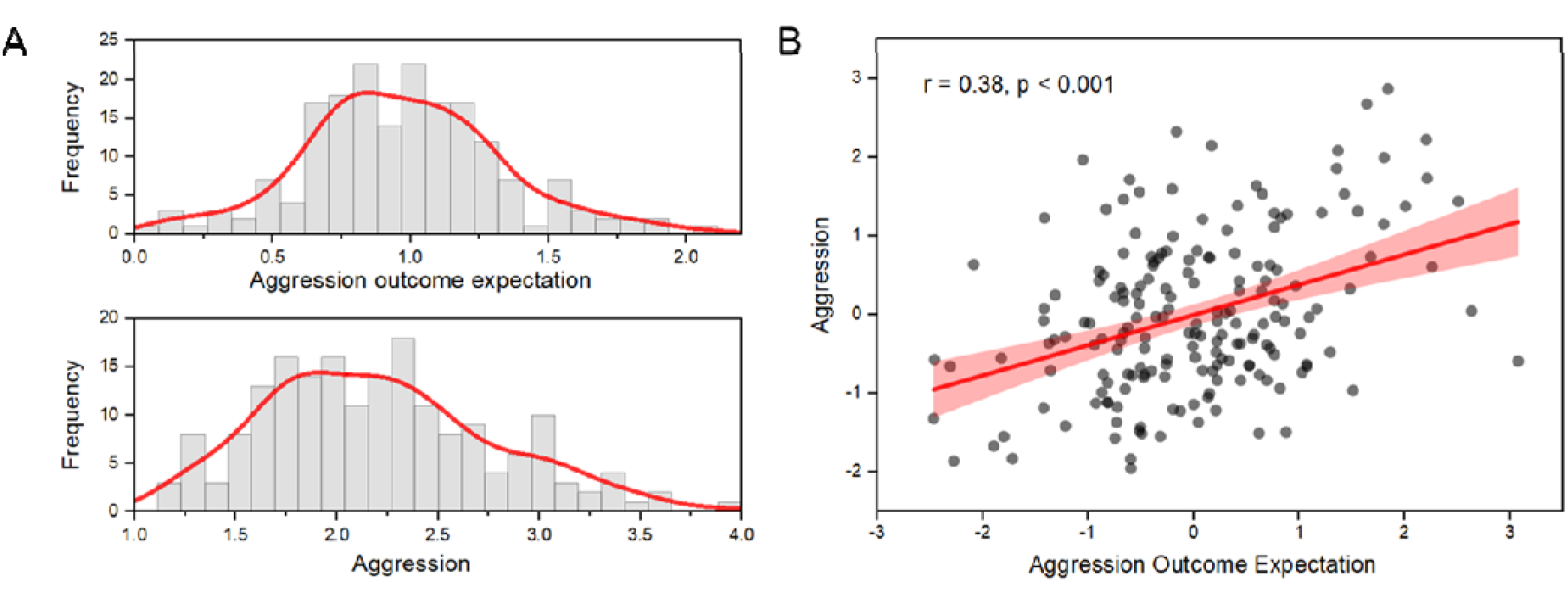
(A) aggression outcome expectation and aggression both follow the normal distribution. (B) The scatter plot showing a positive correlation between aggression outcome expectation and aggression. The values in the plot are standardized residuals after regressing out age, gender, and total intracranial volume.

### sMRI results

To detect underlying brain structures associated with aggression outcome expectation,a whole-brain multiple regression analysis was performed with the GMV. After controlling for age, gender and TIV, aggression outcome expectation was positively correlated with the GMV in the right middle temporal gyrus (right MTG; x = 55.5, y = -58.5, z = 1.5; *t* =3.35; cluster sizes =352, *p* < 0.05, GRF corrected; Figure 2, Table 2). No other significant correlation was observed in the analysis. In addition, to evaluate the stability of the link between the GMV in the right MTG and aggression outcome expectation, we conducted a confirmatory cross-validation analysis. The results revealed that the GMV in the right MTG reliably predicted aggression outcome expectation, *r*_(predicted, observed)_ = 0.23, *p* < 0.001.

**Table 2.**
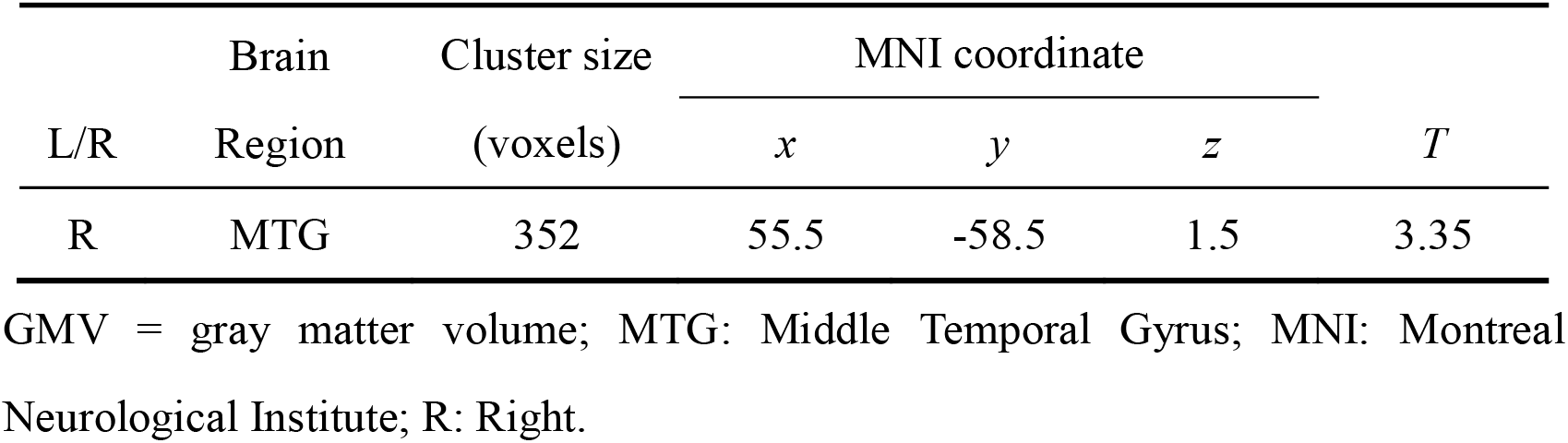
Brain region exhibiting a significant correlation between the GMV and aggression outcome expectation.

**Figure 2.**
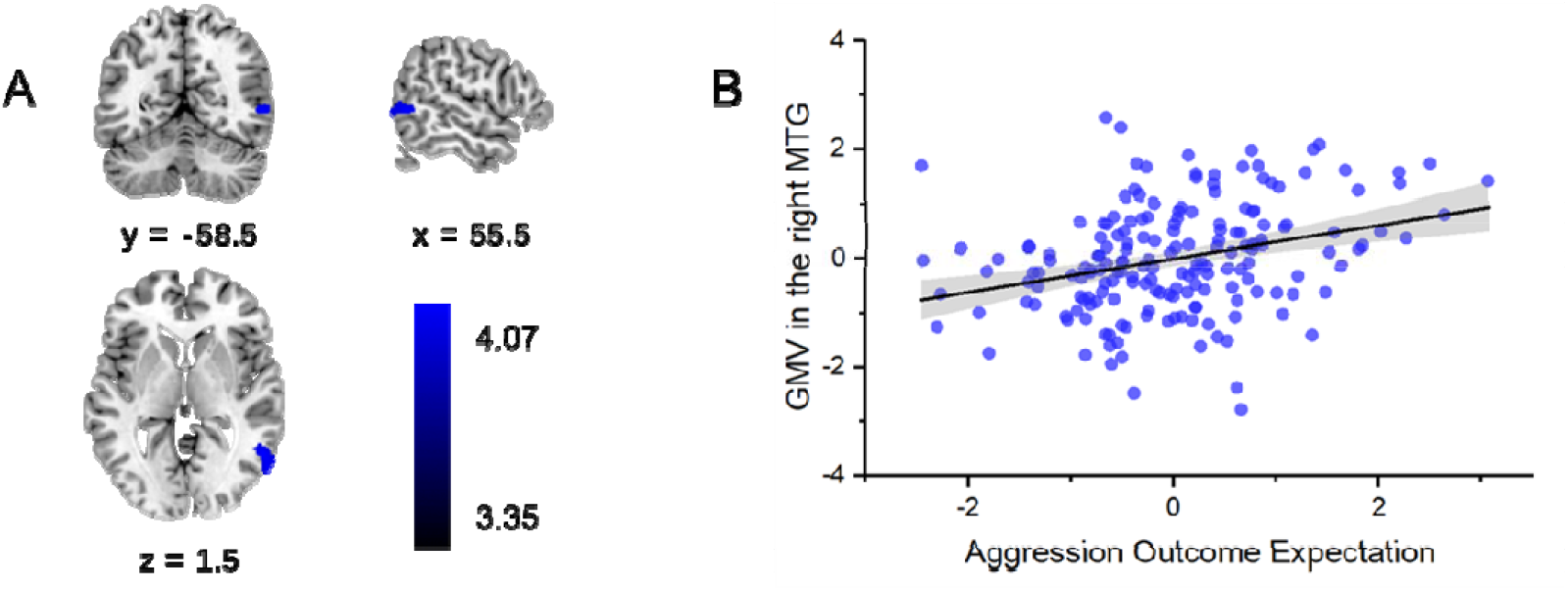
GMV structure correlated with aggression outcome expectation. (A) Brain images depicting the positive correlation between aggression outcome expectation and the GMV in the right MTG. (B) The scatter plot showing the positive correlation between aggression outcome expectation and the GMV in the right MTG after controlling for age, gender, and TIV.

### The mediation model of aggression outcome expectation, aggression and brain structure

In order to explore how aggression outcome expectation affects the relationship between brain structure and aggression, we performed a mediation analysis. The results revealed that aggression outcome expectation played a significant mediating role between the GMV in the right MTG and aggression (indirect effect = 0.17, CI [0.07, 0.28], p < 0.05; Figure 3). There is no significant association between the GMV in the right MTG and aggression, but the influence of GMV in the right MTG on aggression was reduced when was included as a mediator. The relationship between the GMV in the right MTG and aggression was established by aggression outcome expectation.

**Figure 3.**
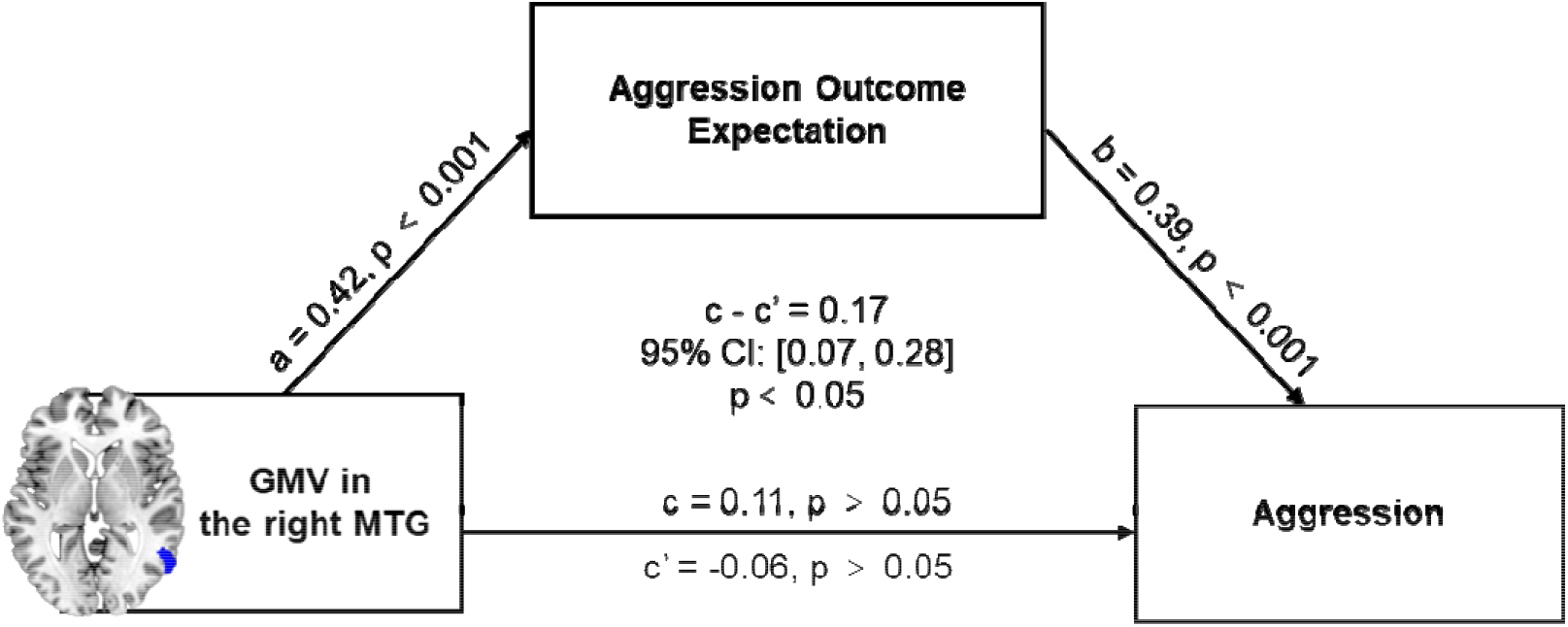
aggression outcome expectation mediates the influence of the GMV in the right MTG on aggression. The graph shows the path diagram of the mediation analysis in which aggression outcome expectation mediates the association between the GMV in the right MTG and aggression. All path coefficients are standard regression coefficients.

In addition, another mediation analysis was also conducted to examined whether brain structure mediated the relationship between aggression outcome expectation and aggression. The results showed that the relationship between aggression outcome expectation and aggression could not be mediated by the GMV in the right MTG (indirect effect = -0.01, CI [-0.06, 0.03], *p* > 0.05).

All mediation models’ analyses controlled for covariates including sex, age, and total intracranial volume (TIV).

## Discussion

The present study aimed to investigate the neural correlates of aggression outcome expectation and its association with aggression using voxel-based morphometry (VBM) analysis. Our findings shed light on the neural mechanisms underlying aggression outcome expectation and its potential impact on aggressive behavior.

The significant positive correlation observed between aggression outcome expectation and regional gray matter volume (GMV) in the right middle temporal gyrus (MTG) provided novel insights into the neural basis of this cognitive factor behind aggression. The right MTG’s involvement suggested its role in processing and integrating information related to anticipated outcomes of aggressive actions. This aligned with previous research highlighting the MTG’s engagement in socio-emotional processing and understanding complex social cues (Wang et al., 2021; Wang et al., 2018; Yang et al., 2016). Our results extended this understanding by implicating the right MTG in aggression outcome expectation, highlighting its importance in the context of aggressive tendencies.

Furthermore, our study revealed that aggression outcome expectation mediated the relationship between GMV in the right MTG and aggression. This suggested that the structural characteristics of the MTG may influence aggression by shaping individuals’ expectations regarding the consequences of their aggressive behaviors. The mediation effect underscored the intricate interplay between neural structure and cognitive processes, indicating that alterations in brain regions like the MTG may exert their influence on behavior through cognitive pathways, such as outcome expectation.

The current findings held implications for the broader understanding of aggression and its neural underpinnings. By identifying the neural correlates of aggression outcome expectation, we contributed to the growing body of knowledge that seeks to unravel the complex interplay between cognition and behavior. Moreover, our study suggested a potential avenue for identifying neurobiological markers associated with aggression. The discovery of specific brain regions, such as the right MTG, as key players in aggression outcome expectation offered a promising direction for future research aiming to develop targeted interventions and preventive strategies for aggression-related issues.

However, it is important to acknowledge certain limitations of our study. The cross-sectional nature of the data limits our ability to establish causality or temporal relationships between neural correlates and aggression outcome expectation. Longitudinal studies could provide further insights into the dynamic changes in brain structure and aggression-related cognitive processes over time. Additionally, the use of self-report measures for aggression and outcome expectation introduces potential biases and warrants cautious interpretation of the results.

In conclusion, this study provided valuable contributions to the field by elucidating the neural underpinnings of aggression outcome expectation and its connection to aggressive behavior. By integrating cognitive and neural perspectives, we advanced our understanding of the complex nature of aggression, offering potential avenues for targeted interventions and the identification of neurobiological markers associated with aggression.

## Data Availability

All data produced in the present study are available upon reasonable request to the authors

## Acknowledgments

We thank all the participants and all research collaborators from Southwest University, Beijing Normal University, Key Laboratory of Cognition and Personality, Ministry of Education, Southwest University.

